# Intraoperative phrenic stimulation offsets diaphragm fiber weakness during cardiothoracic surgery

**DOI:** 10.1101/2022.09.16.22279894

**Authors:** Guilherme Bresciani, Thomas Beaver, A. Daniel Martin, Robbert van der Pijl, Robert Mankowski, Christiaan Leeuwenburgh, Coen A.C. Ottenheijm, Tomas Martin, George Arnaoutakis, Shakeel Ahmed, Vinicius Mariani, Wei Xue, Barbara K. Smith, Leonardo F. Ferreira

**Author notes:** **Corresponding Author:** Leonardo F. Ferreira, University of Florida, 1864 Stadium Rd, 100 FLG, Gainesville, FL 32611-8205. equal contributors (Barbara K. Smith, Leonardo F. Ferreira). **Author Contributions:** Conception of the work – TB, CL, CACO, DM, LF, BS Acquisition of data – GB, TB, RM, DM, TM, GA, SA, BS Analysis and interpretation of the data – GB, TB, RV, CL, CACO, DM, LF, BS Drafting the work – GB, TB, RV, CACO, LF, BS Critical revisions of the work – RM, CL, DM, TM, GA, SA.

## Abstract

**Rationale:** Mechanical ventilation rapidly induces slow and fast fiber contractile dysfunction in the human diaphragm, which could be attenuated by phrenic nerve stimulation. Here, we present data from a controlled trial of intraoperative phrenic stimulation to offset slow and fast fiber contractile dysfunction and myofilament protein derangements.

**Objectives:** In this study, we tested the hypothesis that intraoperative hemidiaphragm stimulation would mitigate slow and fast fiber loss of contractile function in the human diaphragm.

**Methods:** Nineteen adults (9 females, age 59 ±12 years) consented to participate. Unilateral phrenic twitch stimulation was applied for one minute, every 30 minutes during cardiothoracic surgery. Thirty minutes following the last stimulation bout, biopsies were obtained from the hemidiaphragms for single fiber force mechanics and quantitation of thin filament protein abundance. Effects of stimulation and fiber type on force mechanics were evaluated with linear mixed models with the subject treated as a random intercept effect.

**Measurements and Main Results:** Subjects underwent 6 ±2 hemidiaphragm stimulations at 17 ±6 mA, during 278 ±68 minutes of mechanical ventilation. In slow-twitch fibers, cross-sectional area (p<0.0001) and specific force (p<0.0005) were significantly greater on the stimulated side. Longer-duration surgeries were associated with lower slow-twitch specific force (p<0.001). Stimulation did not alter contractile function of fast-twitch fibers or calcium-sensitivity in either fiber type. There were no differences in abundance or phosphorylation of myofilament proteins.

**Conclusion:** Unilateral phrenic stimulation during open chest surgery preserved contractile function of slow-twitch diaphragm fibers, but had no effect on relative abundance of sarcomeric proteins.

## INTRODUCTION

Mechanical ventilation (MV) is life-saving treatment and required for open-chest surgeries. However, extended periods of MV can lead to ventilator-induced diaphragm dysfunction (VIDD), which increases chances of medical complications and has been recognized as a major clinical barrier to weaning among patients who require prolonged MV support (1). The signs of VIDD occur within hours of MV and include increased protein degradation and loss of force generation (2-6). The loss of force in VIDD occurs due to fiber atrophy and contractile dysfunction (4, 7, 8), with the latter being documented by a decrease in force divided per cross-sectional area (specific force). The exact mechanisms of VIDD, especially in humans, are unknown but appear to involve unloading of the mechanosensitive sarcomeric protein titin (9). Titin extends from the Z-disk to the M-band of the sarcomere and contributes to elastic properties of the sarcomere (10). Unloading by MV induces passive shortening of the diaphragm (11), and changes in sarcomere length induce conformational changes in titin, which trigger signaling pathways involved in protein turnover (12, 13). Prolonged controlled MV leads to the degradation of multiple sarcomeric elements within diaphragm fibers, including the primary contractile protein, myosin heavy chain, as well as titin (14). Sarcomeric protein degradation is a putative mechanism for contractile dysfunction in VIDD (13).

Activity-based interventions, such as preoperative inspiratory muscle training, have been proposed as a method to maintain diaphragm loading and prevent VIDD (15). However, pre-MV interventions do not directly treat the inactivity that causes diaphragm dysfunction *during* MV. Diaphragm activation during MV is possible via electrical stimulation of the phrenic nerve, which prevents diaphragm atrophy in animals (16, 17) and humans (18, 19), and it partially protects against contractile dysfunction in rodents during prolonged MV (16). We reported that intermittent intraoperative phrenic stimulation increased force of fast-twitch diaphragm fibers in a small pilot study with a cohort of patients undergoing open-chest surgery (20). Positive outcomes in pre-clinical studies and our earlier pilot findings led us to complete the current study to test the hypothesis that intermittent electrical phrenic stimulation during open-chest surgery protects from sarcomeric protein degradation and diaphragm fiber contractile dysfunction induced by MV.

## METHODS

### Patients

Eligibility criteria for intraoperative hemidiaphragm stimulation included adults (18-80 years) scheduled for non-emergent clinical cardiothoracic surgeries anticipated to last at least 3 hours, enabling 4 or more bouts of phrenic stimulation. Patients were ineligible to participate for any of the following: (1) history of surgery to the diaphragm or pleura, (2) severe obstructive lung disease (FEV1 <40% predicted), (3) restrictive lung disease, (4) severe chronic heart failure (NYHA class IV), (5) chronic kidney disease with serum creatinine >1.6 mg/dL, (6) body mass index <20 or >40 kg/m2, (7) chronic uncontrolled metabolic disease, (8) concurrent neoplastic or myopathic illness, or (9) use of immunosuppressants, corticosteroids, or aminoglycoside antibiotics within 28 days of surgery. The protocol was approved by the University of Florida Institutional Review Board and prospectively registered (NCT03303040). Patients were recruited from the University of Florida cardiothoracic surgery clinic, and each patient provided their written informed consent in advance of participation.

### Pulmonary Function Testing

Subjects completed forced vital capacity (FVC) maneuvers to screen for significant obstructive disease. Seated spirometry (Discovery 2, FutureMed) was conducted in accordance with recommended guidelines (21). Additionally, maximal inspiratory pressure (PImax) was measured with a commercial manometer (MicroRPM, Vyaire Medical) using recommended procedures (22). For both tests, trials were repeated 3-5 times, and the highest effort was reported.

### Intraoperative Phrenic Stimulation

All subjects underwent a midline sternotomy. During their surgery, the right or left phrenic nerve was stimulated using an external cardiac pacemaker (Model #5388, Medtronic) and temporary pacing wires. Selection of the stimulated side was determined by the surgeon, based on ease of access to the corresponding phrenic nerve and the type of procedure being performed. The surgeon placed electrodes adjacent (∽5-10 mm) to the phrenic nerve, either near the pulmonary artery or at the right pericardium, and confirmed the stimulating electrodes did not directly contact the phrenic nerve. Twitch stimulation parameters included a 1.5 ms pulse width, frequency and 1-minute duration. Atrial output was 0 mA, and ventricular output was set at twice the twitch threshold, which ensured a vigorous hemidiaphragm contraction. Stimulations continued every 30 min from the time the phrenic nerves were initially exposed until completion of the procedure.

### Diaphragm Biopsies

Approximately 30 minutes following the final stimulation bout, diaphragm biopsies (∽150 mg) were obtained from the stimulated and unstimulated costal hemidiaphragms and processed as described in Supplemental Methods.

### Statistical analyses

Demographic characteristics of the patients were summarized using n (%) and Mean ± SD. Single fiber outcomes of interest (area, abs force, passive tension, maximal specific force, Ktr, H and KD) were summarized with Mean, SD, and range. Then we summarized the average measures by side and fiber type. The individual fiber data were used to compare effects of stimulation on contractile function via a linear mixed model (SAS 9.4). Relationships between fiber CSA, specific force, and duration of MV were evaluated with linear regression. The abundance of proteins was tested for normality and compared with a Wilcoxon test.

## RESULTS

### Patient Characteristics

**Figure 1** outlines the subject enrollment information. Of 1,018 surgical clinic patients reviewed for eligibility, 25 subjects were enrolled. Three subjects were enrolled but did not receive intraoperative stimulation, due to preoperative changes in their clinical care plan. One subject initiated but did not complete intraoperative stimulation due to the requirement of high-dose neuromuscular blockade as part of surgical care. Of 21 patients who completed intraoperative hemidiaphragm stimulation, single fiber mechanics were not evaluated in two patients, due to unavailability of study personnel (n=1), and secondary to non-cardiac surgery (n=1). **Table 1** illustrates the demographic characteristics of the sample (n=19, 9 female). Patients underwent an average of 6 ±2 intraoperative stimulations with stimulation amplitude of 17 ±6 mA, and they were mechanically ventilated for 284 ±62 minutes prior to biopsy. Of the 38 biopsies obtained for this study, two specimens (subject 15: unstimulated hemidiaphragm; subject 17: stimulated hemidiaphragm) contained primarily fat and connective tissue.

**FIGURE 1.**
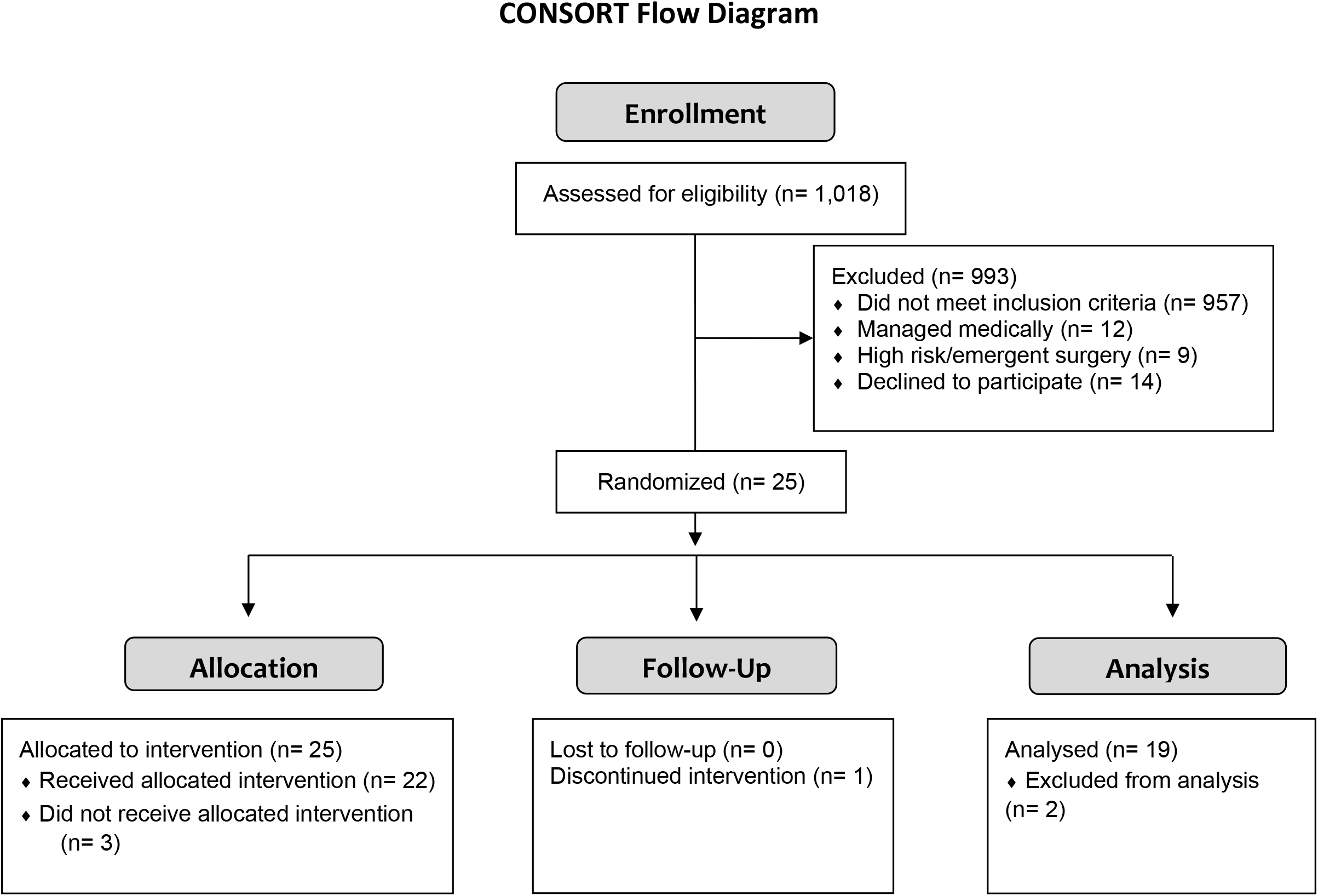
CONSORT diagram.

**Table 1.** Patient characteristics.

|  | Mean (SD) |
| --- | --- |
| Age (years) | 59 (12) |
| gender | 9F/10M |
| BMI (Kg/m<sup>2</sup>) | 30 (4) |
| %FVC | 88 (12) |
| %FEV1 | 85 (13) |
| PI Max (cmH<sub>2</sub>O) | 90 (23) |
| MyHC1 (%) | 64 (10) |
| Time (min)<br>Intubation to biopsy | 278 (68) |

F: female; M: male; BMI: body mass index; %FVC: forced vital capacity; %FEV1: forced expiratory volume in the first second; PImax: Maximal inspiratory pressure; CABG: coronary artery bypass grafting; AVR: aortic valve replacement; MVR: mitral valve replacement; MyHC1: type 1 myosin heavy chain; v: number of vessels grafted

### Diaphragm Single Fiber

In total, 745 diaphragm fibers were activated. Of those, 382 were classified as slow (186 stimulated and 196 unstimulated) and 363 fibers were classified as fast (186 stimulated and 177 unstimulated). **Figures 2, and S2 – S4** show the effects of stimulation on single fiber size and contractile function. In slow fibers, absolute force (**Figure 2A**), cross-sectional area (**Figure 2B**), and specific force (**Figure 2C**) were significantly higher (10-25%) for the stimulated hemidiaphragm (all p<0.005). **Figure 2D** shows specific force vs pCa relationship for data averaged per subject. **Figure S2** shows that there were no significant differences in calcium sensitivity (pCa50), rate-constant of force redevelopment (ktr, cross-bridge kinetics), and Hill coefficient (cooperativity). In fast fibers, we did not detect significant differences between stimulated and non-stimulated hemidiaphragms for any of the variables examined (**Figure 2E-H and S1**). We also examined the relationship between fiber area × force (**Figure S3**) and MV duration × specific force (**Figure S4**) in stimulated and non-stimulated hemidiaphragms. Fiber force was directly related to estimated cross-sectional area in slow (p<0.001) and fast fibers (p<0.001) as expected, with a steeper slope for stimulated slow fibers (p = 0.001) that is consistent with higher specific force (**Figure S3**). Averaged specific force was inversely related to MV duration in unstimulated slow fibers (unstimulated fibers: p<0.005, stimulated fibers: p=0.153,) further supporting protection of VIDD with stimulation (**Figure S4**). There was no significant relationship between force and MV duration in fast fibers (stimulated fibers: p=0.239, unstimulated fibers: p=0.649).

**FIGURE 2.**
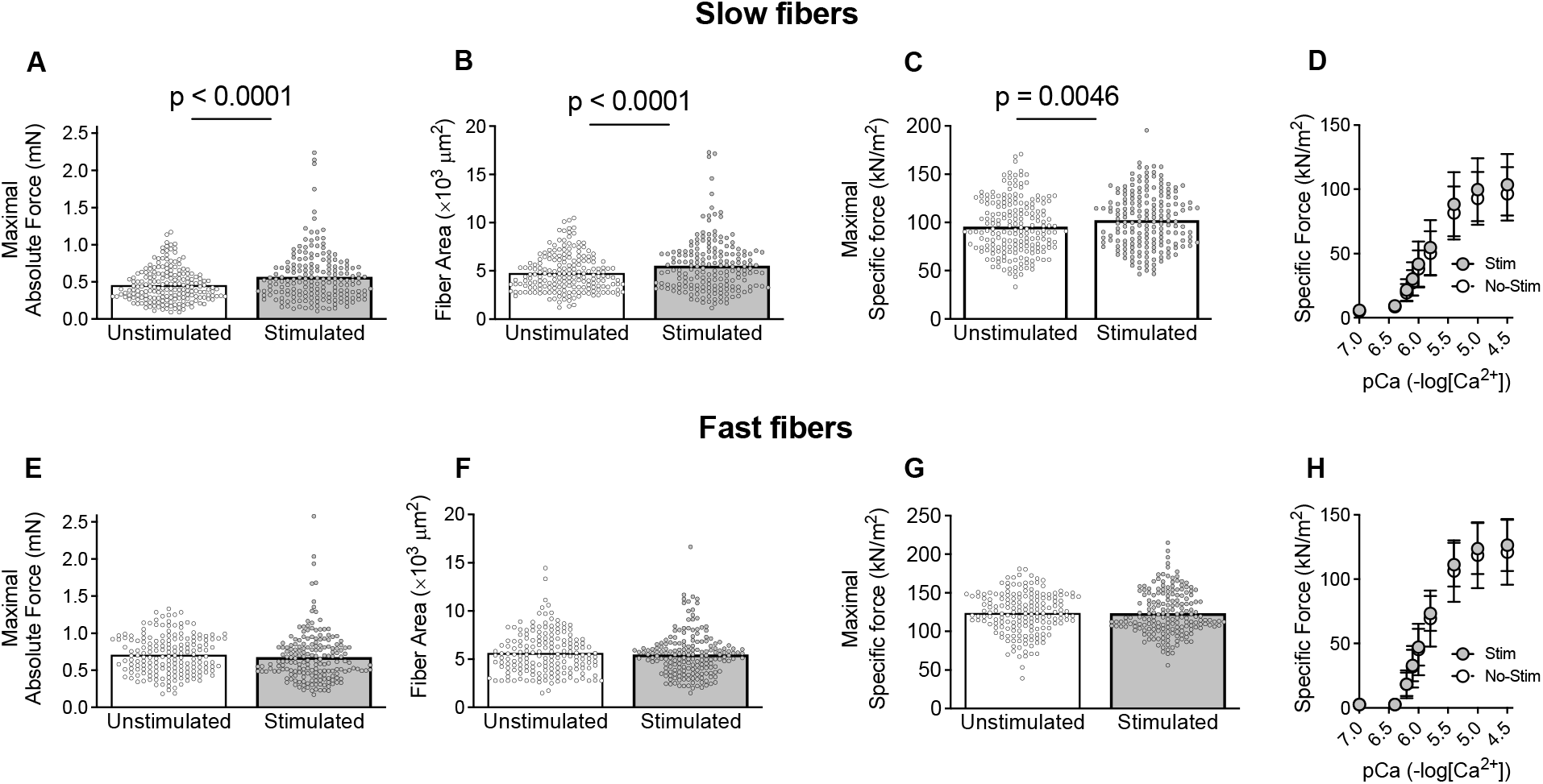
In slow fibers, maximal calcium activated force (**A**), fiber cross-sectional area (**B**), and maximal specific force (**C**) were significantly greater in the stimulated hemidiaphragm. Averaged specific force vs pCa relationship (**D**). In fast fibers, no significant differences in maximal force (**E**), fiber cross-sectional area (**F**), maximal specific force (**G**), or the specific force vs pCa relationship (**H**) were detected. Mixed effect model; mean and standard deviation shown. A-C and E-G dots are data from individual fibers. Statistical analysis by linear mixed model.

### Protein abundance and post-translational modifications

There were no significant differences between sides in the relative abundance of the key structural, regulatory, and contractile proteins titin, nebulin, MyBP-C, MyHC, or actin. Similarly, no significant differences between stimulated and non-stimulated were evident in the abundance of titin or its degradation products (**Figure 3**), titin phosphorylation status (**Figure 4**), or abundance of titin-associated proteins (**Figure 3**). Total phosphorylation levels of nebulin and myosin binding protein C were also not different between sides (**Figure 4**). The protein calpain (CAPN3), which interacts with titin and is involved in sarcomere and cytoskeletal remodeling, has been implicated in VIDD. We measured total CAPN3 and CAPN3 degradation products (markers of activation) and found no difference between sides (**Figure 3B**). Similarly, the levels of the stress response proteins vinculin and CSRP3 (**Figure 3C**) were similar for each side. Altogether the analyses of myofibrillar proteins, phosphorylation, and titin-linked or stress-responsive proteins revealed no differences between stimulated and unstimulated diaphragm.

**Figure 3.**
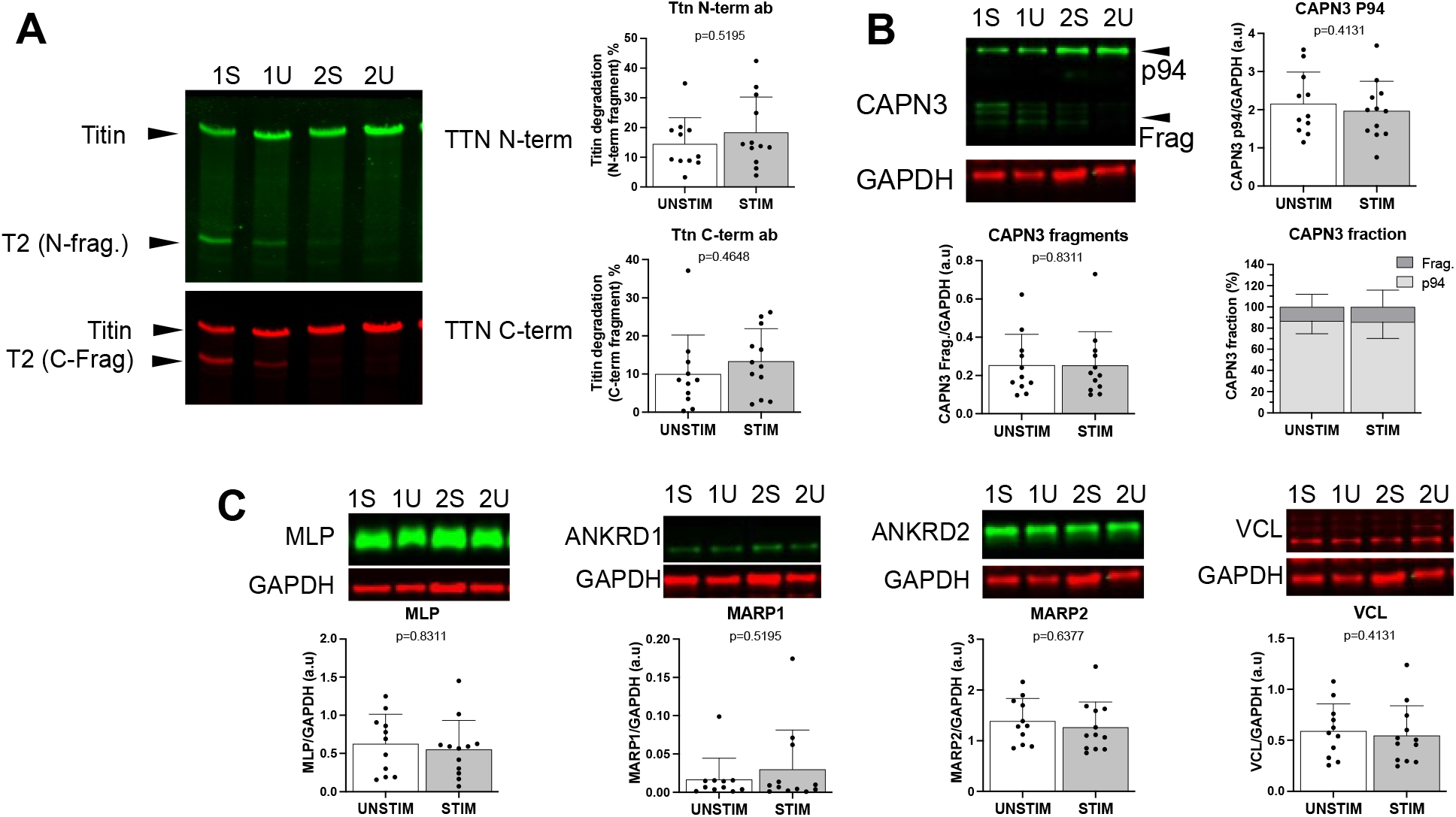
Content of titin fragments and titin-binding proteins. Western blot images and data of the N-terminal and C-terminal fragments of titin (**A**). (**B**) Calpain-3 (CAPN3), muscle specific protease at the N2A region of titin. Full-length, inactive calpain-3 (P94) and calpain fragments comprise the CAPN3 fraction. (**C**) Titin-binding proteins. Muscle Lim-Protein (MLP), telethonin-binding protein in the Z-disk region; muscle ankyrin repeat proteins 1 and 2 (MARP1 and MARP2), stress-responsive proteins at the N2A region of titin; and vinculin (VCL), membrane protein involved in sarcomere stability. Sample Western blot images are from the stimulated (STIM) and unstimulated (UNSTIM) sides of subjects 1 and 2. P-values shown from Wilcoxon tests; mean and standard deviation shown.

**FIGURE 4.**
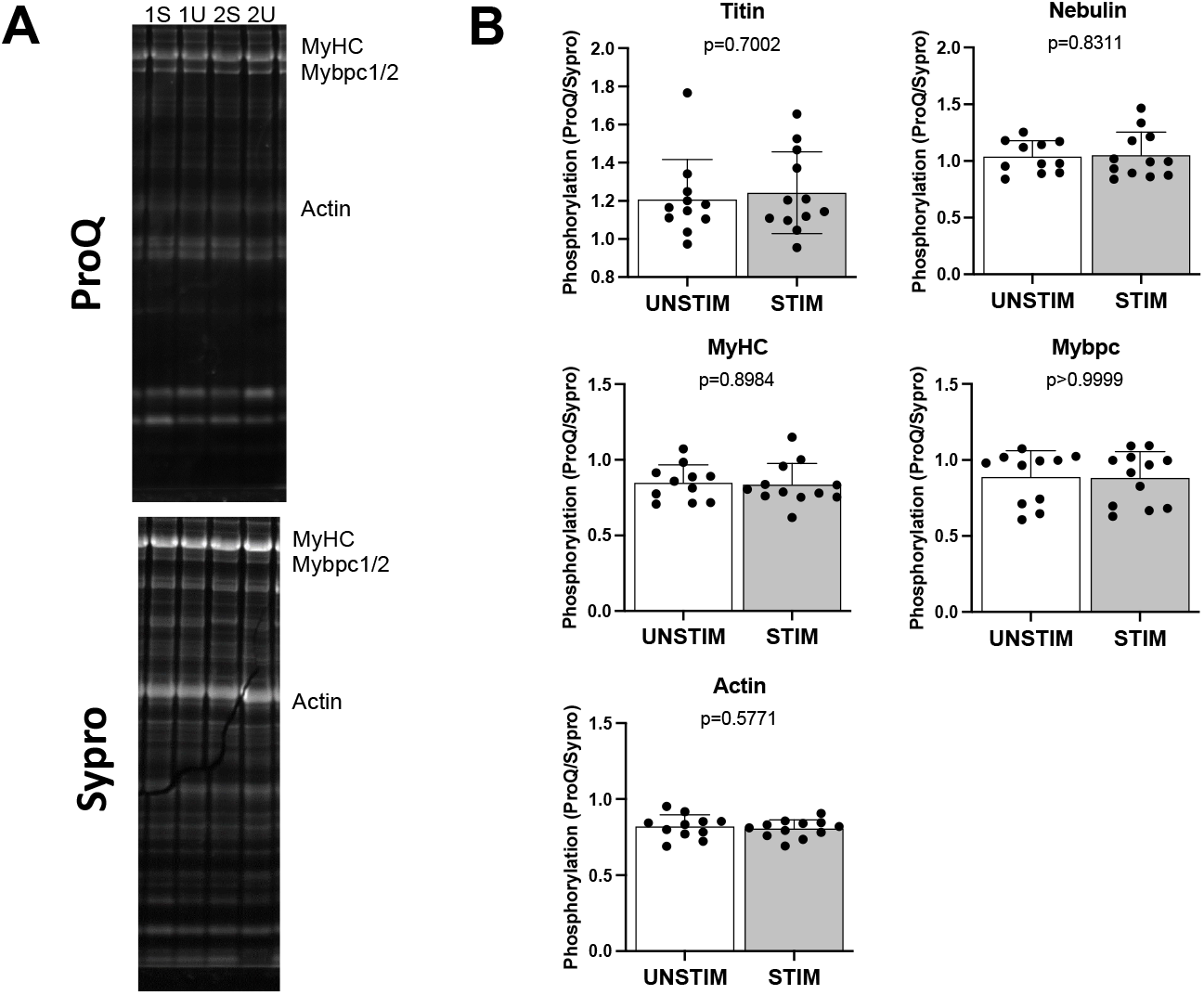
Sarcomere protein phosphorylation. (**A**) Sample images of Pro-Q (phosphorylation) and SYPRO Ruby (total protein). Images are from the stimulated (STIM) and unstimulated (UNSTIM) sides of subjects 1 and 2. (**B**) Figures depict phosphorylation state of titin, nebulin, myosin heavy chain (MyHC), myosin binding protein C (Mybpc), and actin. P-values are from Wilcoxon tests; mean and standard deviation shown.

## DISCUSSION

The primary novel findings of our study were that intermittent phrenic stimulation during 4-6 hours of cardiothoracic surgery resulted in higher hemidiaphragm fiber forces and estimated cross-sectional area in slow fibers. We also found an inverse relationship between non-stimulated slow fiber contractile function and the duration of MV. Fast fibers were not affected by stimulation. The higher force in slow fibers was not accompanied by changes in abundance and phosphorylation status of myofibrillar proteins or titin-linked proteins measured in whole-tissue homogenates.

### Clinical implications and current therapies for VIDD

Findings from this study may have implications for clinical management of patients recovering from cardiothoracic surgery. Sonographic evidence of poor pre-operative diaphragm function is independently associated with post-operative pulmonary complications and increased ICU length of stay (23). Additionally, serial ultrasounds identified post-operative diaphragm dysfunction in 38 of 100 patients, following elective cardiac surgeries (24). While no preoperative patient attributes distinguished those who eventually developed diaphragm dysfunction, average cardiopulmonary bypass time was 87% longer in patients with postoperative diaphragm dysfunction. Post-operative diaphragm dysfunction resulted in more frequent pulmonary complications, leading to slower weaning from MV and longer ICU recovery time (24, 25). A recent large multinational study found that 55% of patients who underwent cardiac surgery with cardiopulmonary bypass experienced at least one post-operative pulmonary complication (26). While several risk factors were identified, included among them were intraoperative PEEP and neuromuscular blockade, which can worsen existing diaphragm dysfunction (27, 28). While our findings suggest potential to attenuate diaphragm fiber contractile dysfunction with intraoperative phrenic stimulation during controlled MV, further studies are needed on its postoperative functional impact.

### Impact of mechanical ventilation on diaphragm fiber size and contractile function

The consensus from animals and human studies is that MV causes diaphragm atrophy and impairs contractile function (4, 7, 29, 30). However, a previous study showed that MV, in the absence of critical illness, did not impair contractile function of permeabilized single fibers (31). While we must be carefult with direct comparisons between studies, these findings could suggest that the loss of pressure generation capacity in humans (32, 33) and contractile function in intact diaphragm bundles (4, 34, 35) after MV result from impairments in neuromuscular transmission, membrane depolarization, and calcium handling. Our study was not designed to establish the effects of MV alone on contractile function. Nonetheless, we found an inverse relationship between slow fiber specific force and time from onset of MV to biopsy. This novel finding suggests that MV causes a progressive decline in sarcomeric protein function in diaphragm fibers of patients who are not critically ill. The contrasting findings between the current and previous study (31) may arise from different modes of ventilation, or exacerbation of VIDD during open-chest surgery (36). During open-chest surgery, the diaphragm is not subjected to the same mechanical forces as when MV occurs with a closed chest (e.g. shortening due to PEEP and cyclic stretches). The surgical procedures performed in our study also required cardiopulmonary bypass, which can further impair diaphragm contractile function (37). Further, protective ventilation during surgery has gained recent consideration (38), but MV modes used during surgery often differ from those used to protect against VIDD in critical illness (39). Overall, studies in humans and animal models support the notion that the diaphragm undergoes rapid (within hours) atrophy and loss of contractile function and, based on our data, open-chest surgery with cardiopulmonary bypass appears to exacerbate these processes. Thus, VIDD may delay weaning of MV and prolong hospital stay not only in patients with critical illness but also those recovering from cardiothoracic surgeries (24).

### Phrenic stimulation during mechanical ventilation

Recent studies suggest stimulation is a feasible, safe, and effective approach to enhance inspiratory pressure production and promote ventilator weaning in patients unable to complete voluntary inspiratory strengthening exercises (40). Studies in animal models and humans have shown that phrenic stimulation during MV prevented or attenuated the decrease in diaphragm thickness, fiber cross-sectional area, and contractile function (16, 17, 19, 41, 42). In comparison to our trial (1Hz stimulation for 60 seconds, every 30 minutes), these previous studies examined longer periods of MV (rats: 18 hours (16), pigs: 19-35 hours (17), humans: 48 to 60 hours (19, 41), sheep: 72 hours (42)) and employed protocols with higher stimulus frequency (20-30 Hz) and train rates (every 20 seconds in rats (16), from every breath to every fourth breath in humans (19, 41), and every breath in sheep (42)). Rat diaphragm specific force assessed in ‘intact’ bundles ex vivo was preserved when MV included phrenic stimulation, but fiber cross-sectional area was not examined (16). In comparison, we observed a higher slow fiber maximal specific force and estimated fiber area in permeabilized fibers in the current study.

The relationship between MV duration and slow fiber specific force suggests that phrenic stimulation did not elicit full protection of sarcomeric protein function. Thus, the greater protection of contractile function in intact rodent diaphragm with phrenic stimulation is most likely due to combined improvements in excitation-contraction coupling and sarcomeric protein function.

Phrenic stimulation during MV in sheep attenuated the decrease in fast fiber cross-sectional area (42). In the current study, slow, but not fast fiber cross-sectional area was greater with stimulation. This discrepancy may be related to differences in stimulation protocols, duration of MV, or species. In pigs, higher diaphragm thickness and fiber cross-sectional area were not accompanied by improved inspiratory pressure (17), and there was no functional assessment reported in humans with higher diaphragm thickness after stimulation during MV (19). Since higher diaphragm thickness did not translate into better function in pigs (17), it is possible that, in larger mammals, phrenic stimulation during MV increases diaphragm thickness partially due to edema, and the higher cross-sectional area may occur through mechanisms that do not preserve contractile function. However, we found that higher slow fiber cross-sectional area in the stimulated hemidiaphragm accompanied proportional increases in maximal fiber force (**Figure S4**). The steeper slope for slow fiber area-to-maximal force relationship in the stimulated hemidiaphragm also reflects the improvement in maximal specific force (**Figure 2**). The slope for fast fibers was also slightly steeper, but not statistically significant (P = 0.054), in the stimulated diaphragm. In general, our study reinforces the safety and feasibility of phrenic stimulation during MV and advances the field by showing intraoperative feasibility and direct benefits to diaphragm fibers.

### Potential mechanisms of VIDD and (partial) protection with phrenic stimulation

MV results in diminished or abolished drive to breathe with a potential for phrenic and neuromuscular transmission dysfunction (43). Unloading the diaphragm triggers disuse atrophy and contractile dysfunction via abnormal proteostasis (increased protein degradation and decreased synthesis), impaired calcium release by the ryanodine receptor 1 (6, 44), and post-translational modifications of sarcomeric proteins (45). Titin-based mechanosensing has been proposed as a signaling hub for diaphragm dysfunction induced by unloading (9). Titin is a critical determinant of myofiber tension at optimal sarcomere length, and phrenic stimulation resulted in higher passive tension. However, there was no detectable effect of phrenic stimulation on titin abundance, degradation products, or titin-binding proteins that modulate stiffness (MARP1 and 2) in whole-tissue homogenates. The greater increase in slow fiber absolute force than specific force suggests that improvements elicited by phrenic stimulation was in proteostasis. Calpains have been implicated in protein degradation, atrophy, and contractile dysfunction induced by MV (46). Yet, we saw no effect of phrenic stimulation on calpain 3 abundance or degradation products that serve as markers of activation. The exact mechanisms underlying sarcomeric protein dysfunction and loss of specific force in VIDD remain unclear (47). The increase in maximal specific force without changes in calcium sensitivity suggests improvements in function of thick filament proteins. We assessed key thick-filament proteins and found no difference in their abundance or phosphorylation status.

Skeletal muscle unloading causes cross-bridges to enter a super-relaxed state that prevents interaction with actin and force generation (48). One plausible scenario is that phrenic stimulation, by imposing intermittent load on the diaphragm during MV, mitigates the loss of specific force by minimizing the transition of cross-bridges to a super-relaxed state. We did not prepare the samples to assess cross-bridge states but this topic is worth investigating in future studies. Overall, we were unable to define potential molecular mechanisms associated with higher fiber area and force induced by phrenic stimulation in whole tissue homogenates. Fiber type-specific post-translational modification proteomics and synchrotron radiation x-ray diffraction will be necessary to resolve potential biochemical and biophysical events underlying the improvements in fiber size and force with phrenic stimulation.

### Limitations and methodological considerations

We stimulated at twice the minimal threshold (up to 25 mA twitches) to achieve supramaximal stimulation. The clinical environment did not permit us to make quantification of force generation, but we could clearly observe vigorous contractions of the stimulated side. The optimal parameters and timing of phrenic stimulation remains unresolved. Our goal was to test a protocol that was considered ‘minimally disruptive’ for the surgical procedures. However, the benefits we report here serve as impetus to optimize phrenic stimulation protocols and examine impact on duration of MV and post-operative pulmonary complications. We acknowledge that selecting the stimulated hemidiaphragm side based on surgeon discretion is not a statistical randomization. In this initial effort, we had to compromise the ideal experimental design to meet the surgical care of the patient. Moreover, supramaximal stimulation and contraction of a hemidiaphragm applies slight lengthening forces upon the contra-lateral unstimulated side. Diaphragm stretches might be enough to blunt the detrimental effects of MV and inactivity (49), and the unstimulated side may not represent a pure “inactivity” control. In this case, our results would underestimate the benefits of intermittent phrenic stimulation. Finally, we measured protein abundance in whole-tissue homogenates to gain insight into potential mechanisms of protection. However, the effects with our protocol seem fiber type-specific, which would require single fiber analysis that are difficult to complete with the amount of tissue available for our study.

### Conclusion

Intermittent, unilateral phrenic stimulation during cardiothoracic surgery led to greater maximal specific force and fiber cross-sectional area in slow fibers of the stimulated hemidiaphragm, without significantly affecting fast diaphragm fiber function. Maximal specific force was positively associated with fiber cross sectional area and inversely related to MV duration. Abundance and post-translational modifications of myofibrillar proteins and titin-linked proteins from whole-tissue homogenates remained unchanged. These findings suggest that protection of maximal specific force occurred by a mechanism independent of effects on sarcomeric protein degradation. Our study supports further examination of intraoperative phrenic stimulation to optimize protocols for increases in fiber size or force and investigation of its potential to decrease postoperative complications.

## Data Availability

All data produced in the present study are contained in the manuscript and available upon reasonable request to the authors

## ACKNOWLEDGEMENTS

We thank the Division of Cardiothoracic Surgery research coordinators with their regulatory assistance. We are grateful for the participation of the patients.

