## Supplemental data for "Intraoperative phrenic stimulation offsets diaphragm fiber weakness during cardiothoracic surgery"

*equal contributors

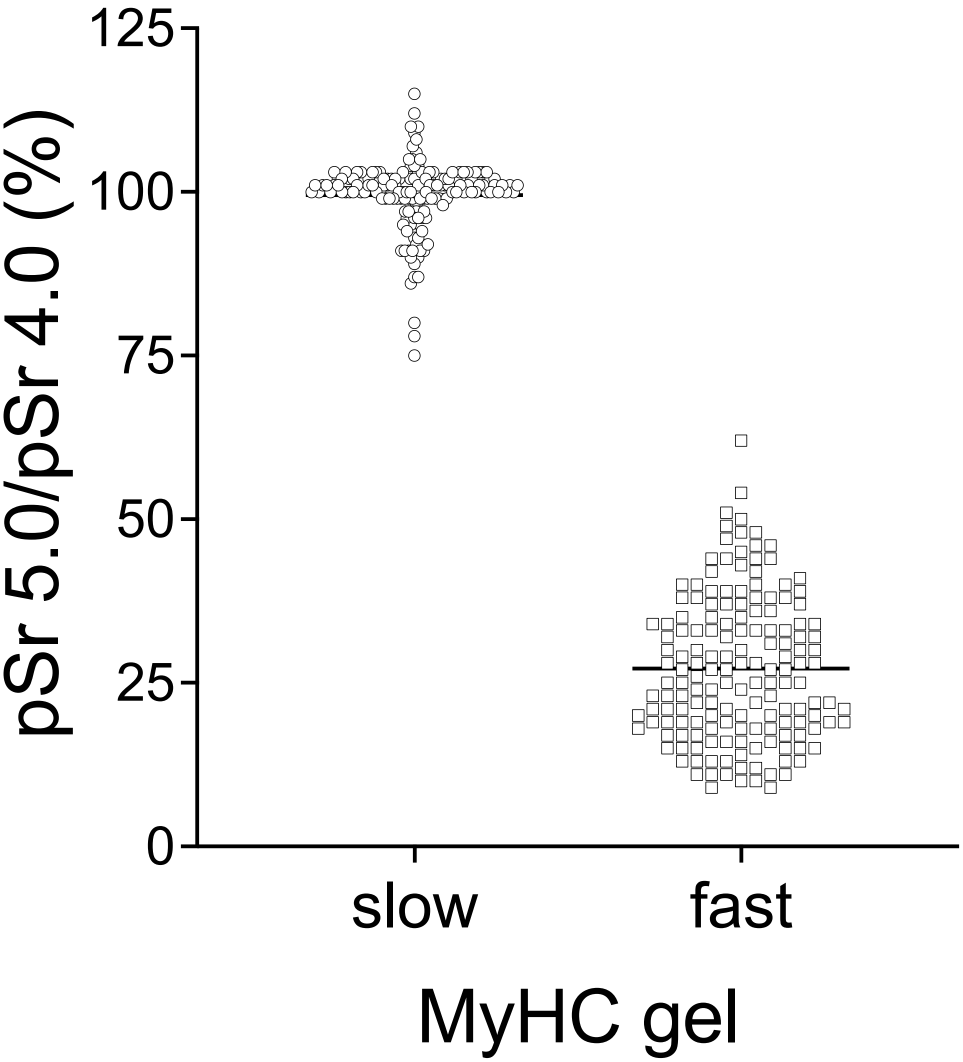


**Figure S1.** Slow and fast myosin heavy chain isoform classification was completed in 345 fibers from 10 subjects, using the ratio between force at pSr 4.0 and 5.0. A ratio of force at pSr 5.0 to pSr 4.0 of 75% or greater indicated slow fibers, and ratio less than 75% indicated fast fibers. The pSr criteria were confirmed with MyHC separation by electrophoresis.

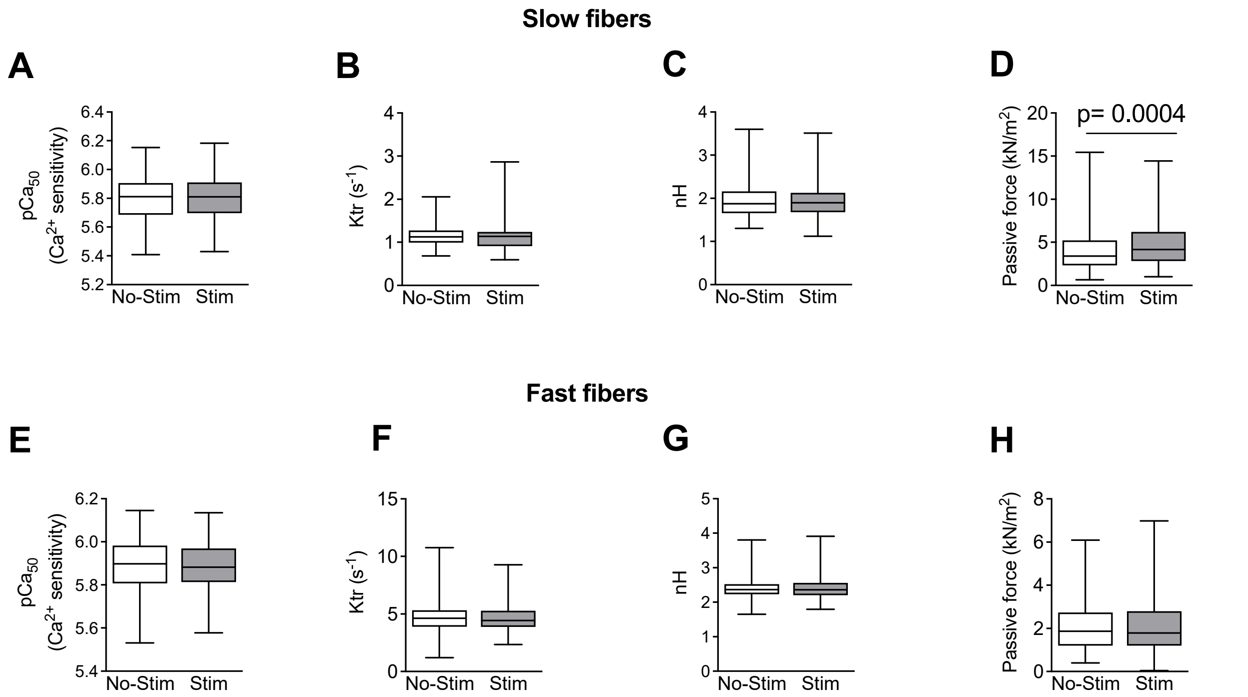


**FIGURE S2.** In slow diaphragm muscle fibers, there were no differences in calcium sensitivity (**A**), rate of tension development (**B**), or the Hill coefficient (**C**). However, maximal passive force (**D**) was significantly greater on the stimulated side of slow fibers. In fast diaphragm muscle fibers, no differences between the stimulated and unstimulated side were found for calcium sensitivity (**E**), rate of tension development (**F**), Hill coefficient (**G**), or passive force (**H**).

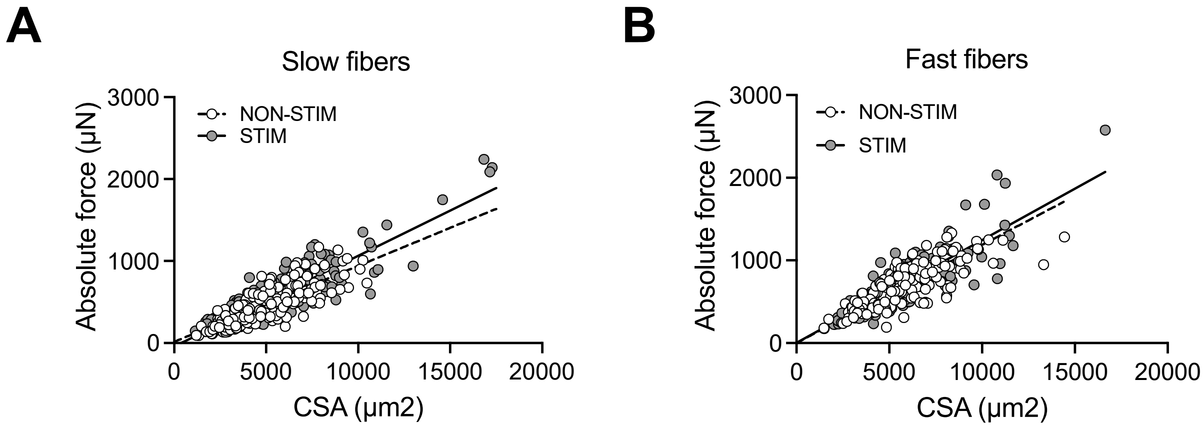


**FIGURE S3. Relationship between fiber cross-sectional area and force.**
In both slow (**A**) and fast (**B**) diaphragm muscle fibers, absolute fiber force was positively associated with the estimated cross-sectional area of the stimulated and unstimulated fibers.

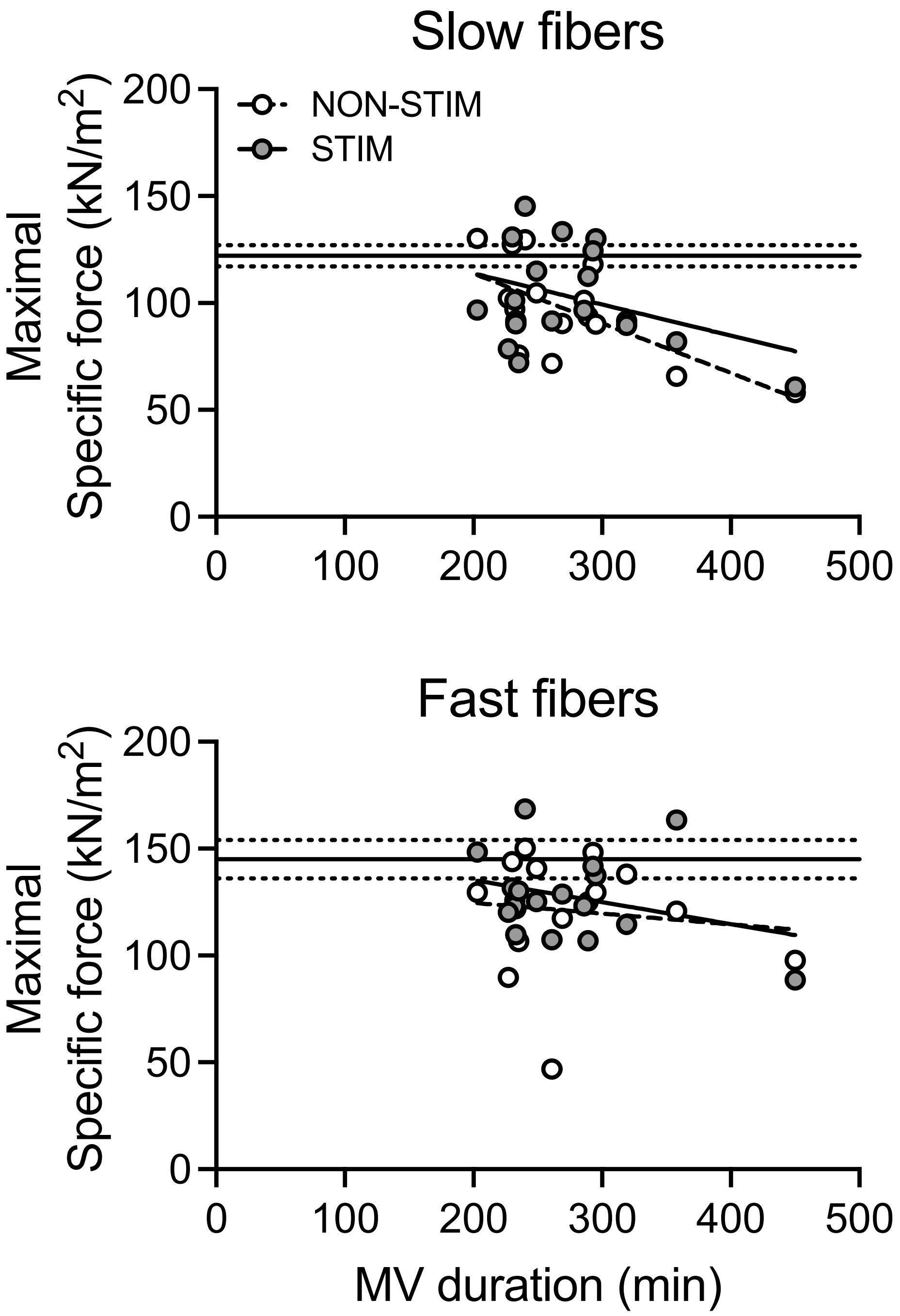


**FIGURE S4.** In non-stimulated (NON-STIM) slow diaphragm fibers, mechanical ventilation (MV) duration was inversely correlated to the maximal specific force. The relationship between specific force and the duration of mechanical ventilation was not significant for fast diaphragm fibers or stimulated slow fibers.
