## supplemental methods for "Intraoperative phrenic stimulation offsets diaphragm fiber weakness during cardiothoracic surgery"

**Diaphragm Biopsies**

Diaphragm biopsy specimens were immediately placed in chilled Buffer X (2.77 mM $CaK_2EGTA$, 7.23 mM $K_2EGTA$, 5.77 mM $Na_2ATP$, 6.56 mM $MgCl_2 \cdot 6H_2O$, 20 mM taurine, 15 mM $Na_2$Phosphocreatine, 20 mM imidazole, 0.5 mM dithiothreitol and 50 mM K-MES, 35 mM KCl; pH adjusted at 0°C to 7.1 using 5 N KOH; 295 mosmol/kg $H_2O$), debrided of extramuscular fat, divided and weighed. Samples specified for single fiber contractile studies were preserved in ice-cold relaxing solution (in mM: 100 KCl, 20 imidazole, 4 ATP, 2 EGTA, 7 MgCl; pH adjusted to 7.0 using KOH), while tissue for titin studies were flash-frozen in liquid nitrogen until further analysis.

**Permeabilized single fiber mechanics**

We prepared samples for permeabilized single fiber mechanics using a technique similar to that described by Campbell and Moss (23), and adapted in our lab for diaphragm muscle (24-26) with a few modifications for this study. Samples were placed in ice-cold relaxing solution and mini bundles (~ 4mm x 1mm) were carefully cut with fine spring scissors to avoid fiber overstretching. Mini-bundles were transferred to relaxing solution with 1% Triton X-100 (skinning solution) for 4 hours at 4°C to achieve chemical permeabilization. Bundles were then transferred to relaxing solution containing 50% glycerol (v/v) for storage at -20°C for up to 3 weeks (23). On the day of the experiment, single fibers were isolated from permeabilized bundles in ice-cold relax solution, then mounted between a force transducer (403B, Aurora Scientific, Ontario, Canada) and a motor arm (312B, Aurora Scientific) in relaxing solution at 15 °C, and stretched to reach a sarcomere length (SL) of ~2.60 μm. Fiber length, width, and height were measured using video microscopy. At this point, we discarded fibers with SL >2.4 μm under slack condition, a sign of damage imposed during fiber isolation in our hands. Fibers with slack SL ≤2.40 μm were then transferred to pCa 9.0 (pCa = - $\log_{10}[Ca^{2+}]$) and allowed ≥3 minutes for equilibration. Calcium-activated force was elicited by immersing the fiber in different pCa solutions at 15°C (in mM: 20 imidazole, 14.5 creatine phosphate, 7 EGTA, 4 MgATP, 1 free $Mg^{2+}$, and free $Ca^{2+}$ ranging from 1 nM [pCa 9.0] to 32 μM [pCa 4.5]) with sufficient KOH (semiconductor grade, Sigma Aldrich) to adjust the ionic strength to 180 mM at pH 7.0. Once force plateaued at each pCa, we performed a quick-release step by rapidly moving the motor arm toward the force transducer by a distance equivalent to 20% of the fiber length, holding for 20 ms, then returning the motor to its original position restoring the initial fiber length. Exposures to each pCa <9.0 were interspersed by a 3-min period at pCa 9.0. This approach minimizes fiber rundown in our hands. Immediately after the last calcium activation, we completed strontium-based activation according to Hvid et al (25) with slight modifications. Briefly, two stock solutions were prepared 1) SrNF (in mM: 90 HEPES, 50 EGTA, 8.5 MgO, 40 $SrCO_3$, 8 $Na_2ATP$ and 10 $Na_2CrP$, pH 7.10 with KOH) and 2) INxF (in mM: 90 HEPES, 50 EGTA, 10.3 MgO, 8 $Na_2ATP$ and 10 $Na_2CrP$, pH 7.10 with KOH), both with free $[Mg^{2+}]$ of 1mM. Solutions INF and SrNF were carefully mixed to yield different pSr concentrations ($-\log_{10}[Sr^{2+}]$): high strontium concentration (pSr 4.0) and intermediate strontium concentration (pSr 5.0). Strontium activation followed the same procedure as described above for calcium activation. We used the ratio between force at pSr 4.0 and 5.0 to define slow and fast myosin heavy chain isoforms, where a ratio of force at pSr 5.0 to pSr 4.0 ≥0.75 indicates slow fibers (slow MyHC) and ratio

<0.75 indicates fast fibers (fast MyHC). Following pSr activation, single fibers were placed individually in dry Eppendorf tubes and stored at -80°C until sample preparation for MyHC isoform analyzes through SDS-PAGE gel electrophoresis. In 10 subjects, we identified 171 slow fibers and 160 fast fibers using pSr. Two bands were identified in 13 fibers (6 from the stimulated side, 7 unstimulated) from 5 subjects. No bands were detected in 1 fiber. We compared the pSr criteria with MyHC separation by electrophoresis for definition of isoforms and found >95% agreement between methods (**Figure S1** and *Supplemental file*). We were unable to obtain pSr results for 5 slow fibers, which were classified using electrophoresis. In four patients, single fibers were classified as slow or fast based solely on the pSr data.

Experiments were performed and analyzed using SLControl software (27). We used fiber width and height to determine fiber cross-sectional area assuming an elliptical shape. The force–pCa relationship of individual fibers was analyzed using a four-parameter Hill equation (Prism 5.0b, GraphPad Software, La Jolla, CA): $F = F_{pas} + F_o(10^{-pCa})^{nH}/[(10^{-pCa})^{nH} + (10^{-pCa50})^{nH}]$, where $F_{pas}$ is passive force, $F_o$ is maximal active force, *n*H is the Hill coefficient, and $pCa_{50}$ is the pCa that elicits half-maximal activation. The force response to the quick-release procedure was fitted using a single exponential equation to determine the rate of tension redevelopment (ktr). All fibers maintained force at pCa 4.5 ≥10% from the first to the last calcium activation.

**SDS-PAGE Analysis of MyHC Isoform**

Single fibers were carefully placed at the bottom of each Eppendorf using a micropipette tip under a microscope. Myosin heavy chain molecules were extracted from each fiber segment by adding 15µl of MyHC extraction buffer containing (in mM) 100 KCl, 100 $KH_2PO_4$, 50 $K_2HPO_4$, 10 EDTA, 16.8 $Na_4P_2O_7$, 4 mM β-mercaptoethanol, 0.5% protease inhibitor cocktail (v/v, P8340, Sigma Aldrich) and pH 6.5 with KOH with 5% Triton X-100 (v/v) according to Tikunov et al (28). Fibers were thoroughly agitated and incubated in shaker for 24 hours at 4°C. After, the final volume was doubled by adding 2x Laemmli Buffer (Bio-Rad) with DTT (0.35M) and samples were heated for 4 min at 95-100°C. At this point, fibers were either directly loaded on gels or stored at -80°C until electrophoresis was performed.

For intact, non-permeabilized whole muscle, frozen diaphragm tissue (-80°C) was used. Approximately 1-2 mg of diaphragm sample were placed immediately in MyHC extraction buffer with 10% Triton X-100 (1:1) and gently ground in ice (1:100 w/v). Samples were then placed in a shaker and incubated for 24h at 4 °C. The following day, 20 µl of the supernatant were placed into a new Eppendorf containing 80 µl of 2x Laemmli buffer (Bio-Rad) with dithiothreitol (DTT; 0.35M) and thoroughly agitated. After adding 30 µl of glycerol per tube, samples were heated (95-100°C) for 4 minutes and then stored at -80 °C until electrophoresis.

We determined the MyHC isoform of single muscle fiber segments and portions of whole tissue from the biopsies. The content of fast and slow MyHC isoforms in single fibers and whole diaphragm samples was determined as previously described by Chung et al (29) with a few modifications. Briefly, the modified version of the protocol consisted in preparing gels in 13.3 x 8.7 cm (W x L) 1.0 mm thick gel casting sets (3459903, Bio-Rad, Hercules, CA). Gels were run at 4 mA (Owl™ EC1000XL, Thermo Scientific, Hempstead, UK) for 36 hours at 4 °C. A diaphragm sample obtained from a Wistar rat was loaded in each gel as an internal standard to assure sample preparation and SDS-PAGE conditions allowed the correct identification of all MyHC isoforms in both single fibers and whole diaphragm samples. Gels were stained using a commercial kit (Silver Stain Plus, Bio-Rad, Hercules, CA) and scanned (Gel Doc EZ Imager; Bio-Rad) for analysis. For single fiber analyzes, we defined fibers as “slow-” or “fast-twitch” and excluded fibers that co-expressed both fast and slow isoforms. For whole diaphragm samples the relative content of each isoform was determined by fitting the densitometry profiles with asymmetric Lorentzian functions (GelBandFitter software) as previously described (30).

**SDS gel electrophoresis and Western blotting for Titin Content**

SDS-PAGE gel electrophoresis and Western blot experiments have been previously described (31). Tissue was ground to a fine powder using Dounce-style homogenizers cooled in liquid nitrogen. Tissue powder was resuspended in a 1:1 mixture of an 8 M Urea buffer (in M; 8 urea, 2 thiourea, 0.05 Tris–HCl, 0.075 DTT, as well as 3% SDS and 0.03% bromophenol blue, pH 6.8) and 50% glycerol containing protease inhibitors (0.04 mM E-64, 0.16 mM leupeptin, and 0.2 mM PMSF). The solutions were mixed for 4 minutes, followed by 10 minutes of incubation at 60°C. Samples were centrifuged at 12.000 rpm and the supernatant was divided into smaller aliquots and flash frozen for storage at −80°C. SDS-PAGE was performed using 1% agarose gels run in a Hoefer SE600X vertical gel system (Hoefer Inc), to separate titin from other proteins. Gels were run at 15 mA per gel for 3 hours. For analysis of titin and MyHC levels gels were stained using Neuhoff's Coomassie brilliant blue staining protocol. Total phosphorylation was analyzed by using Pro-Q Diamond Gel Stain and SYPRO Ruby (Thermo Fisher) for total protein. After staining, gels were scanned using a commercial scanner Gbox (Syngen).

Titin binding proteins MARP1 (ANKRD-1; 1:1000 Myomedix), MARP2 (ANKRD-2; 1:2000 Myomedix) and CAPN3 (CAPN-3; 1:1000 Myomedix), and the stress response proteins CSRP3 (MLP-1; 1:5000 Myomedix) and vinculin (ab18058; 1:2000 Abcam) were quantified using western blot. All proteins were normalized to GAPDH (#2118; 1:5000 Cell Signaling or MA5-15738 1:4000 Pierce). To quantify titin degradation, Western blot was performed using titin N-terminal (H00007273-M06; 1:2000 Abnova) and C-terminal (TTN-9; 1:2000 Myomedix) antibodies. MyHC levels were determined from Coomassie stained initial gels to determine equalized loading for each sample and run on 4-12% acrylamide gels (31). Proteins were transferred onto Immobilon-P PVDF 0.45 μm membranes (Millipore) using semi dry transfer (Bio-Rad). Membranes were blocked with Odyssey blocking buffer (Li-Cor Biosciences) for 1 hour, and subsequently probed with primary antibodies at 4°C overnight. Near Infra-Red dyes were used as secondary antibodies for dual color detection with Odyssey CLx Imaging System (Li-Cor Biosciences).

**Statistical analyses**

Demographic characteristics of the patients were summarized using n (%) and Mean ± SD. Single fiber outcomes of interest (area, abs force, passive tension, maximal specific force, Ktr, H and KD) were summarized with Mean, SD, and range. Then we summarized the average measures by side and fiber type. The individual fiber data were used to compare effects of stimulation on contractile function via a linear mixed model (SAS 9.4), which accounted for the varying number of fibers contrained with each subject's paired stimulated and control samples. Fixed effects included stimulation and fiber type, while the subject was treated as a random intercept effect.

Relationships between fiber CSA, specific force, and duration of MV were evaluated with linear regression. As each patient can have multiple (repeated) observations on each side for each fiber type, the average of the repeated measures for each patient on each side for each fiber type was first calculated to establish relationships with MV duration.

To evaluate the abundance of titin, titin degradation products, and titin-binding proteins, the normality of each dataset was assessed with a Shapiro-Wilk test, and differences between the stimulated and unstimulated sides were compared with Wilcoxon tests. Significance was $p<0.05$.
